# Brain cerebral blood flow with MRI-visible enlarged perivascular space in adults

**DOI:** 10.1101/2024.08.12.24311906

**Authors:** Chunyan Yu, Baijie Wang, Qiyuan Sun, Huiyan Huo, Lingyan Zhang, Hongyan Du

## Abstract

**Objective:** To explore the correlation between enlarged perivascular spaces (EPVS) in the basal ganglia (BG-EPVS) and centrum semiovale (CSO-EPVS) and changes in adult brain cerebral blood flow (CBF).

**Methods:** This cross-sectional single-center cohort study included individuals with varying degrees of EPVS, divided into the BG and CSO based on the established rating scale. Subsequently, the arterial spin labeling (ASL) sequence and its post-processing operation were utilized to obtain CBF values for different grades of BG-EPVS and CSO-EPVS. Logistic regression was conducted to identify risk factors associated with BG-EPVS and CSO-EPVS, and correlation analysis was employed to explore the associations between different grades of BG-EPVS and CSO-EPVS with CBF of the whole brain and specific regions of interest.

**Results:** The regression analysis revealed that BG-EPVS was associated with age (odds ratio [OR]: 1.10, 95% confidence interval [CI]: 1.04–1.15), hypertension (4.91,1.55–15.6), and periventricular white matter hyperintensities (PVWMH) (4.34,1.46–12.95). Conversely, CSO-EPVS was linked to hypertension (4.40,1.43– 13.57), drinking history (2.84,1.08–7.45), sleep duration (2.01,1.19–3.40), and PVWMH (12.20,3.83–38.85). Correlation analysis revealed a negative correlation between BG-EPVS and the CBF of the whole brain (r=-0.28, p=0.00) and most brain regions, except for the brain stem (r=-0.19, p=0.05). Conversely, CSO-EPVS was negatively correlated with CBF of temporal lobe white matter (r=-0.25, p=0.01); however, the significance was lost after FDR correction. CSO-EPVS was not correlated with CBF across various brain regions.

**Conclusion:** Brain CBF decreased with the increasing severity of BG-EPVS, suggesting that BG-EPVS could serve as an imaging marker for reflecting the changes in brain CBF and an effective indicator for early ischemic stroke.

## Introduction

Perivascular spaces (PVS), also known as Virchow-Robin spaces, are fluid-filled spaces around small arteries, capillaries, and venules in the brain parenchyma, containing interstitial fluid and forming a network of excreting channels that remove normal fluid and metabolic waste from the brain.^1^ This is a crucial structure for microvasculature, inflammation, immunodetection, and neuroinflammation, ensuring proper maintenance and a stable neuronal environment.^2^ PVS are considered pathologic when sufficiently enlarged to be visible on magnetic resonance imaging (MRI).^3^ For example, enlarged PVS, or enlarged perivascular spaces (EPVS), may appear in all age groups and are visualized clearly on T2-weighted brain MRI.^4,5^ These EPVS reflect glymphatic stasis secondary to the perivascular accumulation of brain debris, although they may also represent an adaptive mechanism of the glymphatic system to clear them.^6,7^ EPVS are most commonly observed in the regions of the centrum semiovale and the basal ganglia;^8^ still, they have also been identified in the hippocampal regions and frontal cortex. Nevertheless, the exact mechanisms underlying the correlation of EPVS in adults are not fully understood.

Some studies have suggested that EPVS is one of the imaging manifestations of cerebral small vessel disease (CSVD).^9^ In earlier studies, age,^10^ hypertension^11^, and white matter hyperintensities (WMH)^12^ have been identified as risk factors for EPVS in basal ganglia. Also, EPVS has been related to various age-related diseases and neurodegenerative diseases, including brain injury,^13^ Alzheimer’s disease,^14^ Parkinson’s disease,^15^ multiple sclerosis,^16^ stroke or transient ischemic attack (TIA) ,^17^ microbleeds,^18^ and cognitive impairment.^19^ Moreover, studies have suggested that EPVS may reflect the dysfunction of clearance of cerebrospinal fluid and metabolites around cerebral small vessels and microvascular dysfunction.^20^ Interestingly, it has also been discovered that EPVS at different locations in the brain may have different pathological mechanisms. E.g., EPVS in the basal ganglia (BG-EPVS) has been associated with cerebral atrophy in stroke patients and atherosclerosis,^10, 21^ while cerebrovascular amyloid deposition, resulting from impaired interstitial fluid drainage has been associated with EPVS in centrum semiovale (CSO-EPVS).^22,23^ In addition, studies predominantly comprising patients with a TIA/ischemic stroke indicated that BG-EPVS but not CSO-EPVS are prognostic markers of stroke and death, independent of other neuroimaging markers of small vessel disease.^24^ Also, patients with BG-EPVSs were found to be at an increased risk of recurrent ischemic stroke.^25^ However, there is a lack of relevant research investigating the alterations in CBF associated with EPVS.

This study aimed to quantitatively assess the relationship between EPVS and brain CBF, obtained using arterial spin labeling sequences (ASL), which is a non-invasive and cost-effective MRI technique for brain perfusion measurements.^5^ We also investigated the potential effects of different levels of EPVS on brain CBF and explored the possibility of using EPVS as an early imaging marker of changes in brain CBF.

## Materials and Methods

### Patients

Patients treated at Longgang Center Hospital of Shenzhen between March 2023 and May 2024 were recruited in the study. The inclusion criteria comprised those > 18 years old who have undergone cranial MRI and displayed different degrees of EPVS, and their images met the diagnostic quality criteria. Exclusion criteria comprised severe stenosis or occlusion of cerebral blood vessels, concurrent other vascular diseases, history of traumatic brain injury, history of cerebral infarction, history of cerebral hemorrhage, and other diseases or mental disorders that could cause cerebral perfusion or metabolic abnormalities. Also, data with poor image quality were excluded from the analysis.

The Ethics Committee of the Longgang Central Hospital of Shenzhen approved this cross-sectional study. All participants provided written informed consent.

### Demographic and clinical data

We recorded basic demographic data (age, sex, height, weight, alcohol consumption, and smoking) as well as medical and treatment data (hypertension, diabetes, hyperlipidemia, coronary heart disease). Sleep time was recorded as < 6 hours, 6-8 hours, and > 8 hours. Corresponding info was also obtained from medical imaging data.

### MRI Data Acquisition

MR imaging was acquired on the 3.0 T Prsima Siemens scanner. The parameters were as follows: T1 weight imaging (T1WI): TR=250ms, TE=2.5ms, thickness=5mm, FOV =220×220mm², acquisition matrix=320×288; T2 weight imaging (T2WI): TR=4000ms, TE=94ms, thickness=5mm, field of view (FOV)=220×220mm², acquisition matrix=320 × 320; fluid-attenuated inversion recovery (FLAIR): TR=8000 ms, TE=84 ms, thickness=5.0 mm, FOV=220×220 mm², acquisition matrix=320×224; Diffusion weight imaging (DWI): TR=3300ms, TE=54 ms, thickness=5 mm, FOV=220×220 mm², acquisition matrix=160×160; ASL: TR=4600 ms, TE=16.1 ms, thickness=3.0 mm, FOV=220×220 mm², Reconstruction matrix=64×64, post labeling delay = 2000 ms; T1-MPRAGE (high-resolution T1WI for evaluating brain structure) are as follows: TR =2300ms, TE=2.2ms, thickness=1.0 mm, FOV=256×256 mm, acquisition matrix 256×256.

### EPVS assessment

EPVS were defined as cerebrospinal fluid-like signal intensity (hypointense on T1 and hyperintense on T2) lesions in areas supplied by perforating arteries. Those parallel to the imaging plane appeared round and ovoid, and those perpendicular to it appeared linear; their maximum diameter was 3 mm. EPVS were counted in the BG and CSO using the following rating scale: grade 0 = 0 EPVS, grade 1 = 1–10 EPVS, grade 2 = 11–20 EPVS, grade 3 = 21–40 EPVS, and grade 4 => 40 EPVS in each brain region, see Figure 1. For the BG, we excluded perforated substances at the level and below the anterior commissure, as most people have EPVS in this area.^26^ The numbers referred to EPVS on one side of the brain: after reviewing all relevant slices for the assessed anatomical area, the slice and side with the highest number of EPVS were recorded.^27^

**Figure 1.**
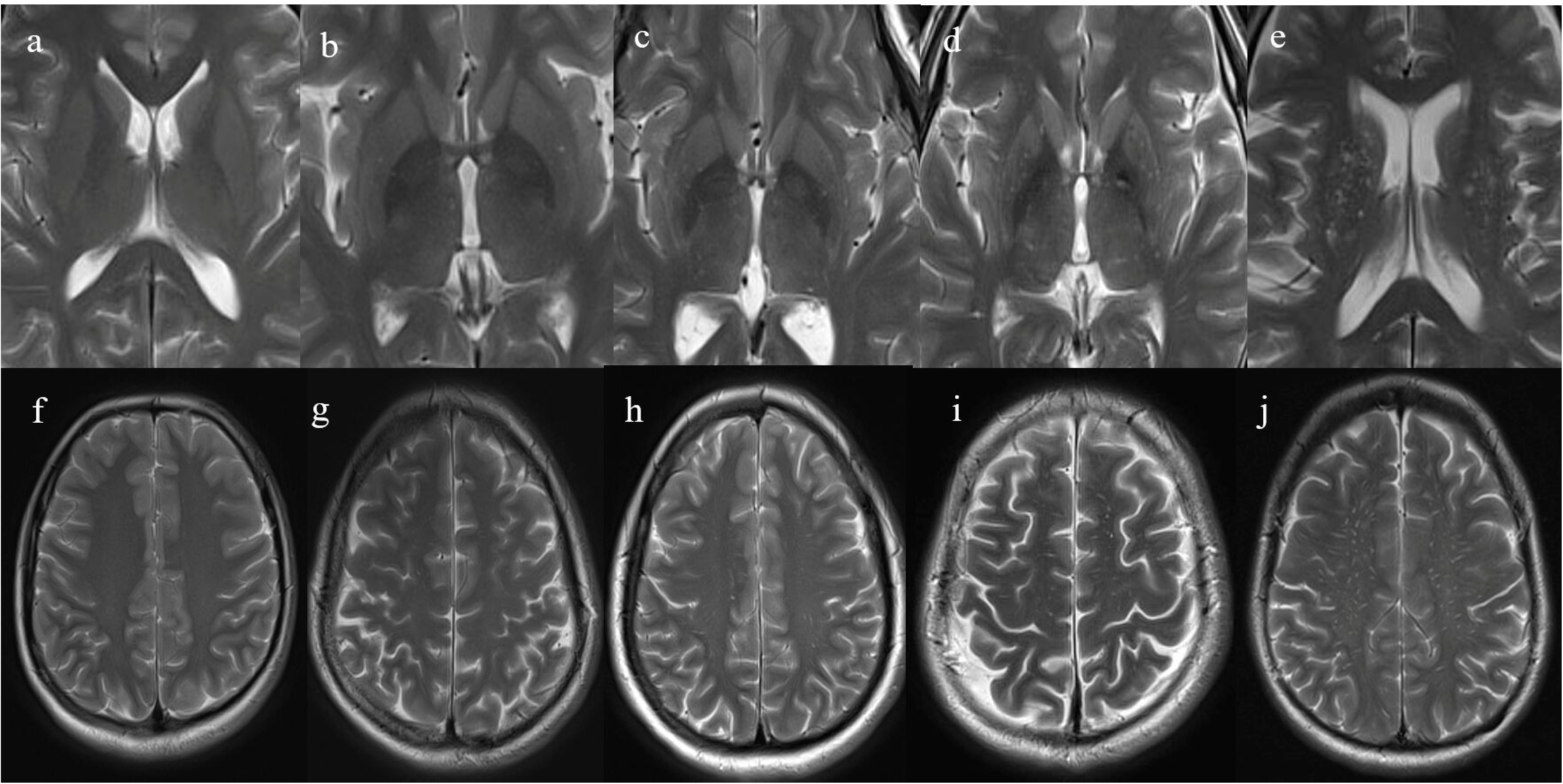
Representative MR images of EPVS burden across severity categories. Panels (a) to (e) show axial T2 MR brain images at the level of the basal ganglia. Panels (f) to (j) show axial T2 MR brain images at the level of the centrum semiovale. Arrows denote individual EPVS. Columns are ordered left to right in increasing bins of EPVS burden (0,1–10, 11–20, 21–40, 41+).

Two trained physicians with > 5 years of experience evaluated the EPVS grade.

Both neurologists discussed any inconsistencies between the reported results until a consensus was reached.

### Fazekas Scale for White Matter Hyperintensities

Fazekas scale was used for white matter hyperintensities (WMH) in both periventricular (PVWMH) and deep white matter (DWMH).^28^ PVWMH was graded as follows: 0 = absence,1 = caps or pencil-thin lining, 2 = smooth halo, and 3 = irregular. DWMH were rated as follows: 0 = absence, 1 = punctate foci, 2 = beginning confluence of foci, and 3 = large confluent areas.

### Extraction of CBF and Segmentation

Figure 2. ASL data were preprocessed and analyzed using Matlab (SPM 12) software. The main analysis steps were as follows: (1) the labeling map of the ASL image was registered to 3D T1 images; (2) 3D T1 images were segmented by NewSegment and registered to the standard Montreal Neurological Institute (MNI) space; (3) CBF obtained from ASL was written into MNI space according to T1 image registration parameters. The CBF values of the whole brain and local brain regions were extracted based on ROI. The gray matter volume (GMV), white matter volume (WMV), cerebrospinal fluid (CSF) volume, and corresponding volume proportions were simultaneously obtained during the segmentation stage.

**Figure 2.**
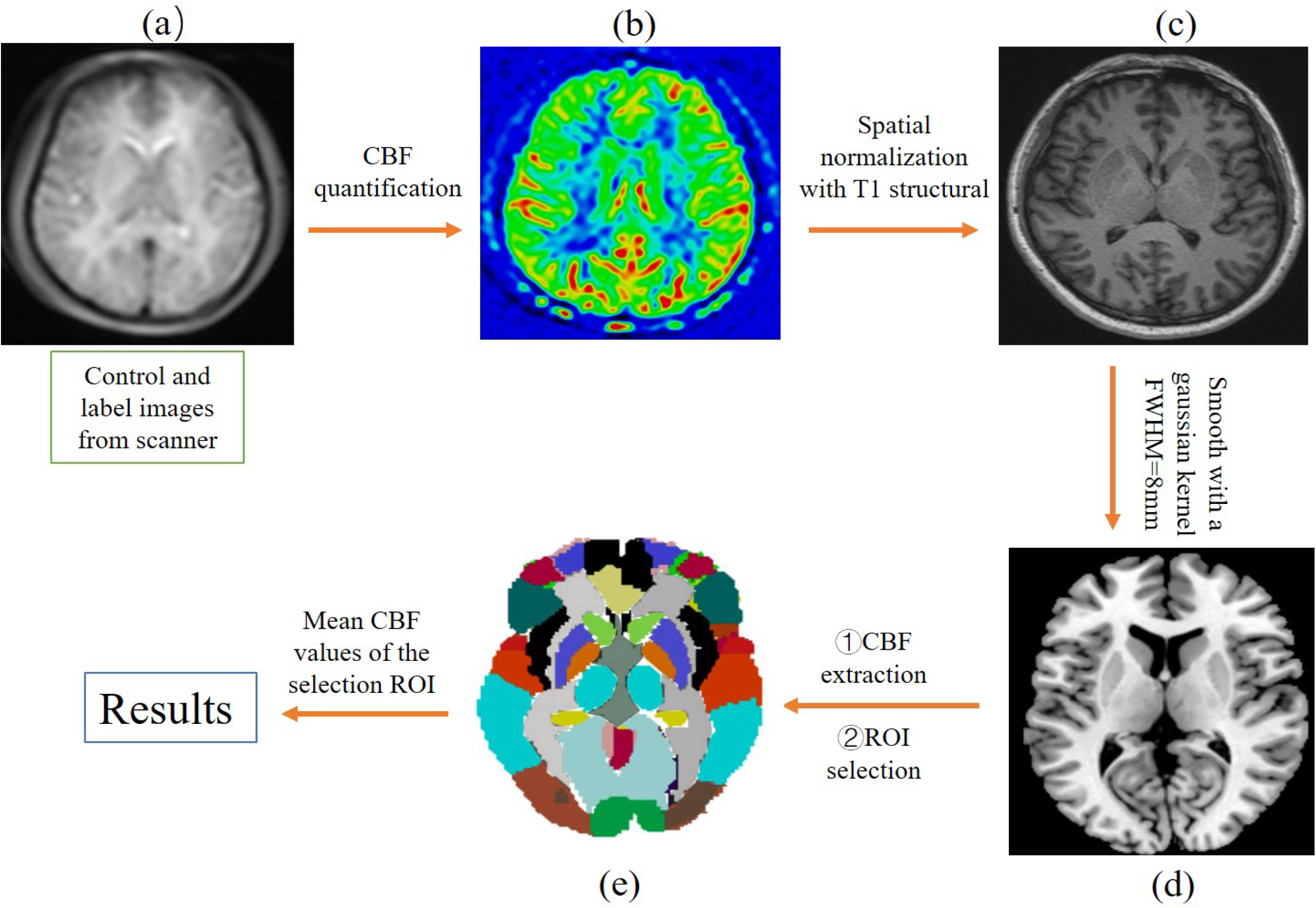
The schematic diagram of the process of extracting cerebral blood flow of brain region interest ((a)-(e)). Note that the images do not represent the corresponding results displayed.

### Statistical analysis

Statistical Package for Social Sciences (SPSS version 26) was used for statistical analyses. Categorical and continuous variables with non-normal distribution were summarized as counts (percentage) and the means (standard deviation, SD) or medians (interquartile ranges, IQR), respectively. Logistic regression was conducted to identify risk factors associated with BG-EPVS and CSO-EPVS. Correlation analysis was employed to explore the associations between different grades of BG-EPVS and CSO-EPVS with CBF of the whole brain and specific regions of interest. The correlation analysis was adjusted for age, gender, hypertension, diabetes, hyperlipidemia, coronary heart disease, smoking, drinking, sleep duration, WMH, and brain volume. p values of correlation analysis were corrected using the false discovery rate (FDR) method with a significance threshold set at 0.05.

## Results

### Participants characteristics

A total of 109 individuals, 55 men and 54 women, with a mean age of 47.1±12.2, were included in the analysis. Participants’ characteristics are summarized in Table 1. Regarding the different BG-EPVS grades, there were 15 cases of grade 0, 22 cases of grade 1, 29 cases of grade 2, 28 cases of grade 3, and 15 cases of grade 4, respectively. Concerning the severity of CSO-EPVS, there were 15 cases of grade 0, 26 cases of grade 1, 34 cases of grade 2, 21 cases of grade 3, and 13 cases of grade 4.

**Table 1.** Patient characteristics.

|  |  |
| --- | --- |
| Demographic data | N=109 |
| Age, mean (SD) | 47.1±12.2 |
| Sex, male, n (%) | 55(50.5) |
| History of hypertension, n (%) | 26(23.9) |
| Diabetes, n (%) | 11(10.2) |
| Hyperlipidemia, n (%) | 17(15.6) |
| Coronary heart disease, n (%) | 4(3.7) |
| Smoking history, n (%) | 21(19.3) |
| Drinking history, n (%) | 28(25.7) |
| BMI, mean (SD) | 23.8±3.7 |
| Sleep duration(hours), n (%) |  |
| <6h | 36(33.0) |
| 6-8h | 41(37.6) |
| >8h | 32(29.4) |
| BG-EPVS, n (%) |  |
| 0 | 15(13.8) |
| 1 | 22(20.2) |
| 2 | 29(26.6) |
| 3 | 28(25.7) |
| 4 | 15(13.8) |
| CSO-EPVS, n (%) |  |
| 0 | 15(13.8) |
| 1 | 26(23.9) |
| 2 | 34(31.2) |
| 3 | 21(19.3) |
| 4 | 13(11.1) |
| PVWMH grade, n (%) |  |
| 0 | 61(56.0) |
| 1 | 34(31.2) |
| 2 | 12(11.0) |
| 3 | 2(1.8) |
| DWMH grade, n (%) |  |
| 0 | 55(50.5) |
| 1 | 41(37.6) |
| 2 | 10(9.2) |
| 3 | 3(2.8) |
| GMV, mean (SD) | 661.6±64.3 |
| WMV, mean (SD) | 493.3±71.9 |
| CSF, median (IQR) | 222.0(195.5-250.0) |
| TIV mean (SD) | 1384.9±145.7 |
| GMVf, median (IQR) | 0.48(0.46-0.50) |
| WMVf, median (IQR) | 0.36(0.34-0.37) |

Note: PVWMH: Periventricular white matter hyperintensities; DWMH: Deep white matter hyperintensities. GMV: Gray matter volume, WMV: White matter volume, CSF: cerebrospinal fluid, TIV: Total intracranial volume, GMVf: Gray matter volume fraction, WMVf: White matter volume fraction, SD: standard deviation, IQR: interquartile range.

### Factors associated with BG-EPVS and CSO-EPVS

Table 2 presents the results of the multivariate analysis comparing clinical and neuroimaging features between BG-EPVS and CSO-EPVS. Regression analysis revealed that BG-EPVS was associated with factors such as age ((odds ratio [OR]: 1.10, 95% confidence interval [CI]: 1.04–1.15), hypertension (4.91,1.55–15.6), and PVWMH (4.34,1.46–12.95). Conversely, CSO-EPVS was linked to hypertension (4.40,1.43–13.57), drinking history (2.84,1.08–7.45), sleep duration (2.01,1.19–3.40), and PVWMH (12.20,3.83–38.85).

**Table 2.** Multivariable ordinal logistic regression analysis showing predictors of increased basal ganglia and centrum semiovale EPVS severity.

|  | BG-EPVS |  | CSO-EPVS |  |
| --- | --- | --- | --- | --- |
|  | OR (95% CI) | P value | OR (95% CI) | P value |
| Age | 1.10(1.04-1.15) | 0.00 | 1.03(0.98-1.08) | 0.21 |
| Sex, Male | 0.58(0.19-1.76) | 0.33 | 0.40(0.13-1.22) | 0.11 |
| History of hypertension | 4.91(1.55-15.6) | 0.01 | 4.40(1.43-13.57) | 0.01 |
| Diabetes | 0.75(0.18-3.16) | 0.70 | 0.50(0.12-1.98) | 0.32 |
| Coronary heart disease | 4.63(0.32-66.2) | 0.26 | 3.99(0.39-40.95) | 0.25 |
| Hyperlipidemia | 0.47(0.14-1.52) | 0.21 | 0.96(0.31-3.02) | 0.95 |
| Smoking history | 1.10(0.39-3.13) | 0.85 | 1.44(0.51-4.07) | 0.49 |
| Drinking history | 2.65(1.00-6.99) | 0.05 | 2.84(1.08-7.45) | 0.03 |
| BMI | 1.09(0.96-1.23) | 0.19 | 1.01(0.89-1.14) | 0.92 |
| Sleep duration(hours) | 1.11(0.66-1.87) | 0.69 | 2.01(1.19-3.40) | 0.01 |
| PVWMH | 4.34(1.46-12.95) | 0.01 | 12.20(3.83-38.85) | 0.00 |
| DWMH | 1.47(0.53-4.03) | 0.46 | 1.15(0.42-3.26) | 0.78 |
| GMV | 1.72(0.80-3.67) | 0.16 | 1.32(0.62-2.76) | 0.47 |
| WMV | 1.71(0.80-3.65) | 0.17 | 1.3(0.61-2.76) | 0.49 |
| CSF | 1.69(0.79-3.58) | 0.18 | 1.21(0.57-2.60) | 0.62 |
| TIV | 0.58(0.27-1.24) | 0.16 | 0.78(0.36-1.64) | 0.50 |

Note: PVWMH: Periventricular white matter hyperintensities; DWMH: Deep white matter hyperintensities. GMV: Gray matter volume, WMV: White matter volume, CSF: cerebrospinal fluid, TIV: Total intracranial volume, GMVf: Gray matter volume fraction, WMVf: White matter volume fraction.

### Association between EPVS and CBF of different brain regions

The CBF values of the whole brain and various brain regions in BG-EPVS and CSO- EPVS are presented in Figure 3 and Figure 4, respectively. As shown in Figure 3, as the severity of BG-EPVS increased, there was a downward trend in the CBF values in specific brain regions. However, with the increasing severity of CSO-EPVS showed in Figure 4, no significant trend in the changes was observed in some brain regions. The specific CBF values of each brain regions with different degrees of EPVS are shown in S1 and S2.

**Figure 3.**
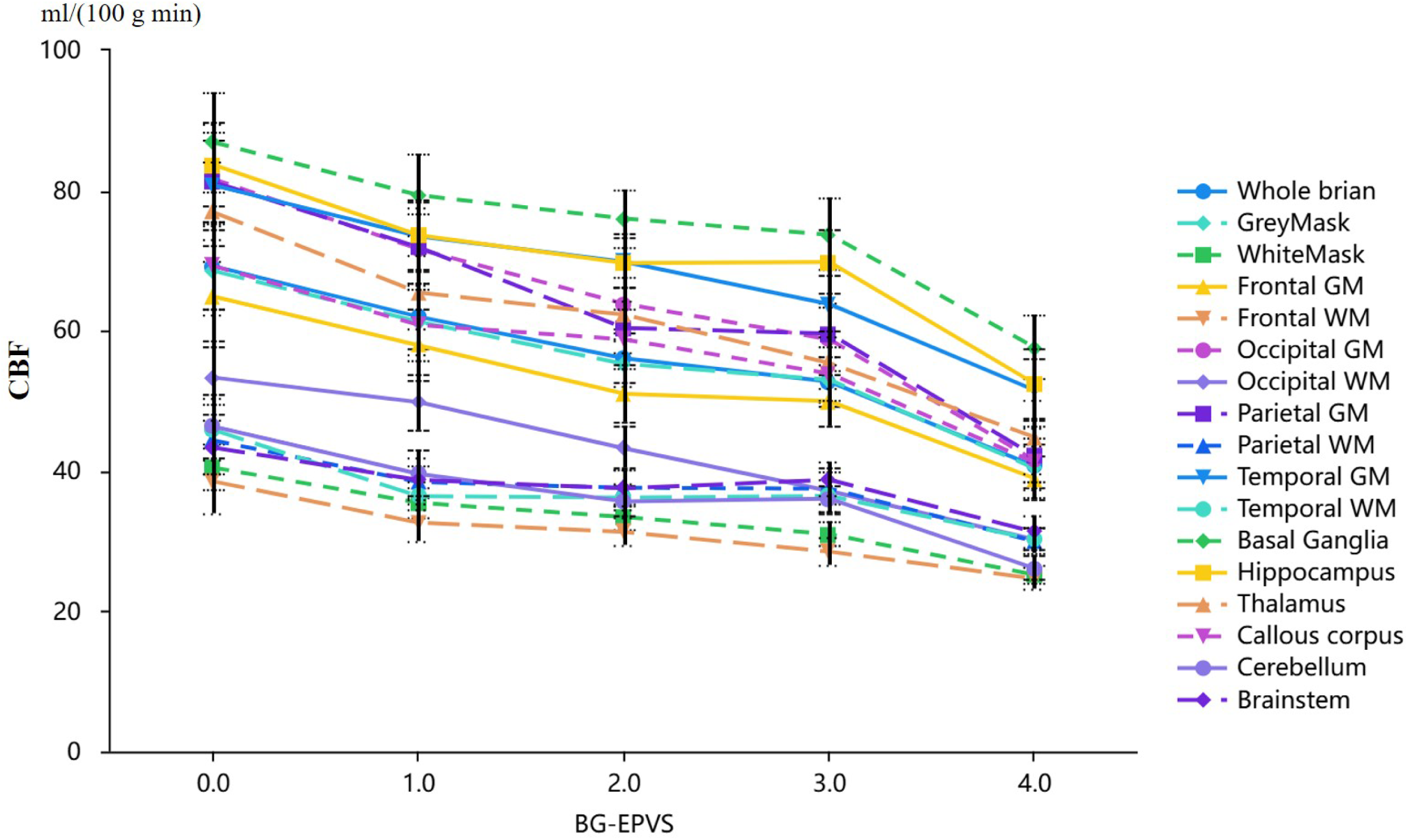
The variations of cerebral blood flow (CBF) across brain regions with varying degrees of BG-EPVS. The x-axis represented the BG-EPVS grade, and the y-axis denoted the CBF values for each specific brain region. Icons (such as triangles and squares) represented the mean CBF, and the dashed lines above and below represented the standard error of the mean CBF.

**Figure 4.**
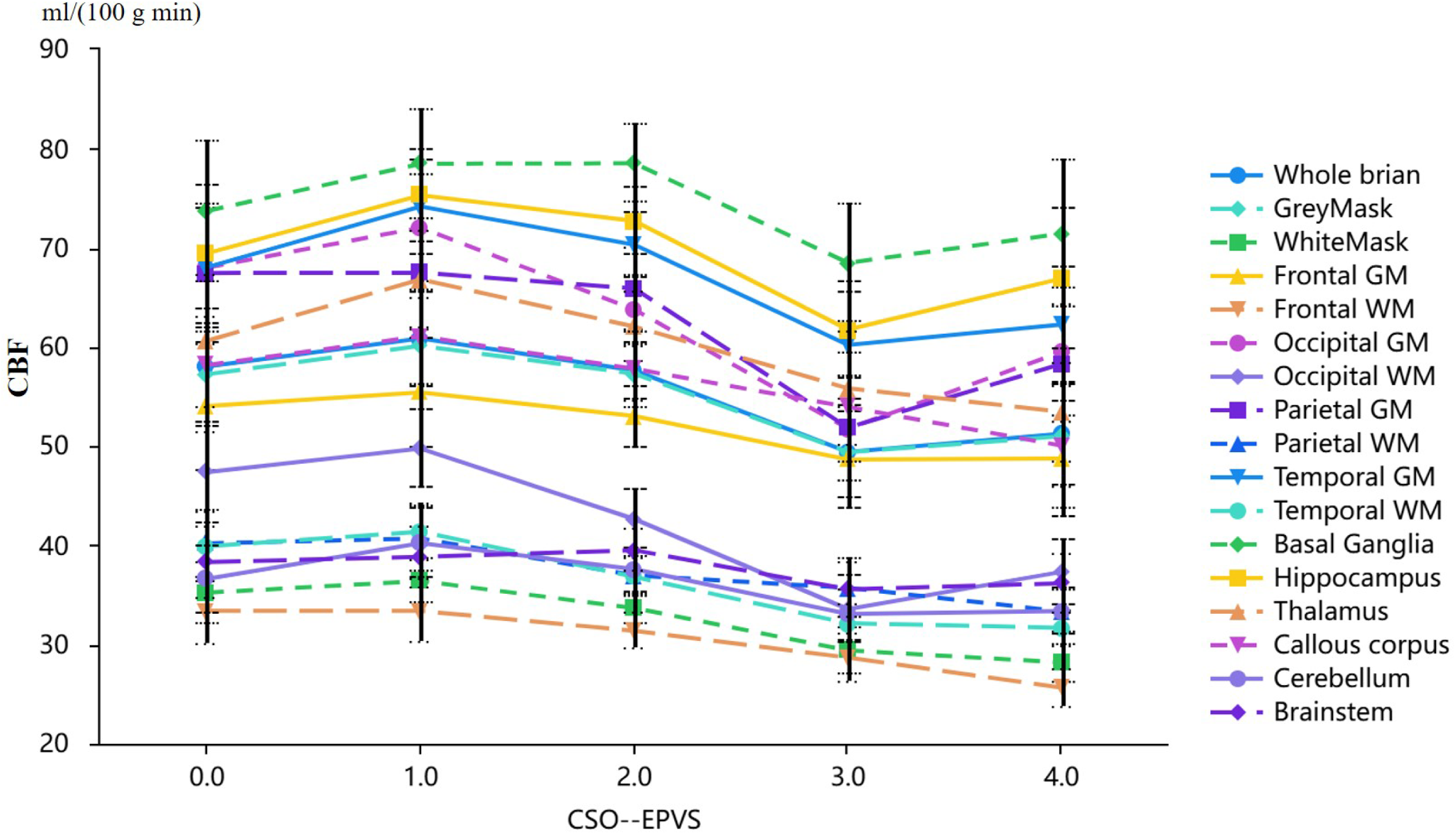
The variations of cerebral blood flow (CBF) across brain regions with varying degrees of CSO-EPVS. The x-axis represented the CSO-EPVS grade, and the y-axis denoted the CBF values for each specific brain region. Icons (such as triangles and squares) represented the mean CBF, and the dashed lines above and below represented the standard error of the mean CBF.

The distribution of P-values corrected with FDR is shown in Figure 5. Correlation analysis revealed that BG-EPVS was negatively correlated with the whole brain (r=- 0.28, p=0.00) and most brain regions, except for the brain stem (r=-0.19, p=0.05). On the other hand, CSO-EPVS showed a negative correlation with temporal lobe white matter (r=-0.25, p=0.01); however, this result was no longer significant after FDR correction. Furthermore, CSO-EPVS showed no correlation with CBF across various brain regions. Unadjusted p values are presented in S3 and S4.

**Figure 5.**
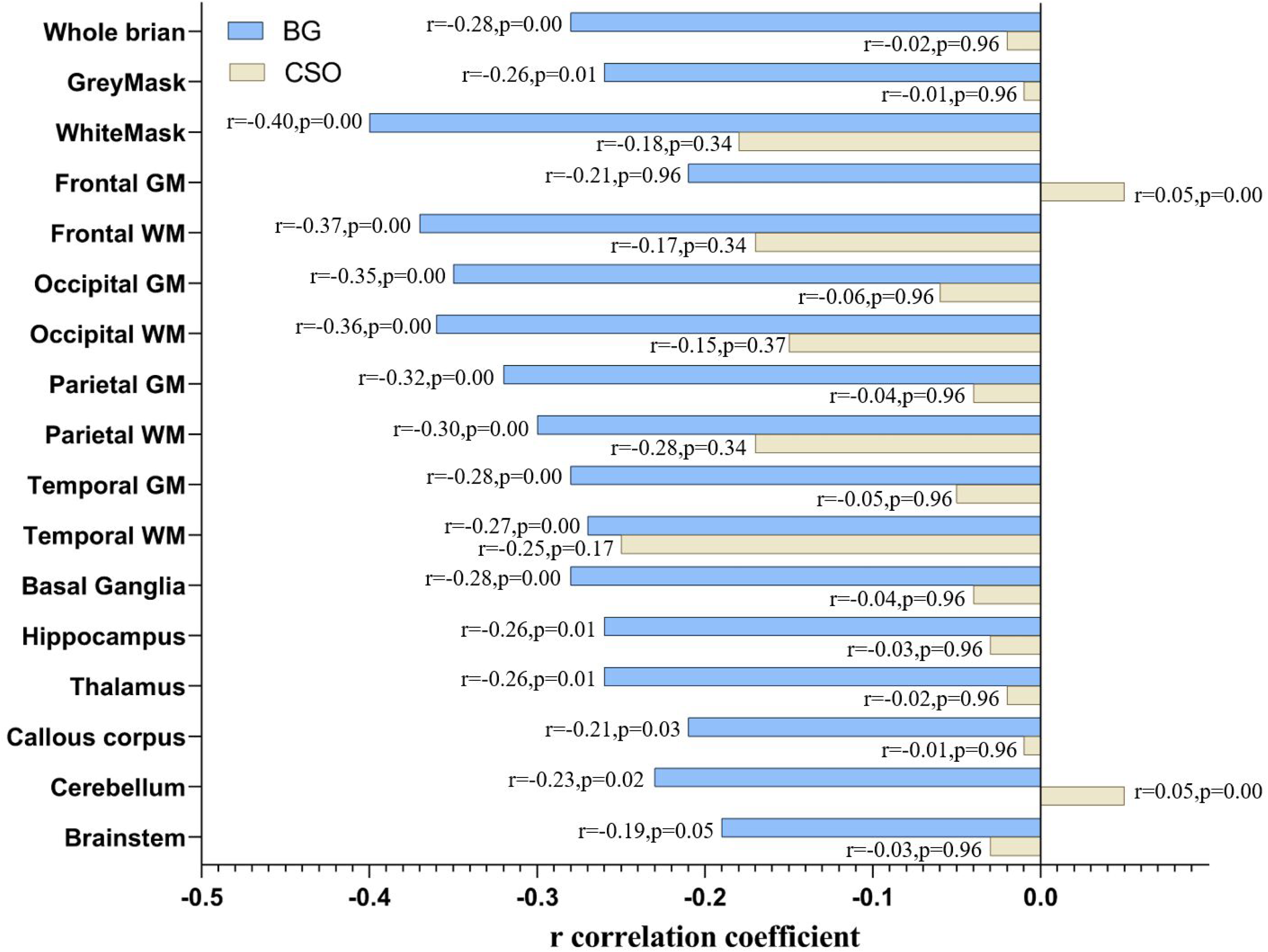
The magnitude of the r correlation coefficient and p values between the different severity of EPVS located in the basal ganglia and semioval center and brain regional cerebral blood flow.

## Discussion

EPVS are most commonly observed in the regions of the centrum semiovale (CSO) and the basal ganglia (BG). In the present study, we quantitatively evaluated CBF changes at different levels of BG-EPVS and CSO-EPVS. Our results revealed that as the severity of BG-EPVS increased, there was a decrease in CBF of the whole brain and almost all regions, which showed a negative correlation, whereas no such phenomenon was observed with CSO-EPVS following the adjustment for multiple factors. In the whole cohort, we identified different risk factors for various locations of EPVS. Age, hypertension, and PVWMH resulted as risk factors for BG-EPVS, while hypertension, smoking, sleep duration, and PVWMH were associated with CSO-EPVS. Our results revealed the correlation between varying degrees of EPVS and changes in brain CBF. The BG-EPVS, rather than CSO EPVS, induced corresponding alterations in brain CBF. Therefore, BG-EPVS could be a potential imaging marker for monitoring CBF and an early imaging indicator for ischemic stroke.

Evidence indicates that the PVS eliminates soluble waste from the brain.^29, 30^ The pathologic mechanism of PVS is not yet fully understood, and current studies have highlighted the important role of aquaporin-4 (AQP4)-lined PVS, cerebrovascular pulsation, and metabolite clearance in normal central nervous system physiology.^20,31^ Our results show the effective removal of waste products in the brain, including Aβ and tau, that accumulate in late-onset Alzheimer’s disease, thereby potentially accelerating vascular Aβ accumulation and perpetuating a detrimental cycle of perivascular clearance dysfunction.^32,33^ The microstructural anatomy, MRI enhancement, physiology, and fluid dynamics of PVS are crucial in neurobiology.

Nevertheless, certain studies indicate that BG-EPVS and CSO-EPVS may be associated with distinct pathological mechanisms.^34^

We observed that as the severity of BG-EPVS increased, a decrease in brain CBF occurred, leading us to speculate that the primary pathogenesis may be associated with atherosclerotic disease. Previous literature reported that BG-EPVS is an imaging biomarker associated with hypertensive arterial disease and arterial stiffness.^35^ Arterial stiffness is a characteristic feature of aging that may contribute to the EPVS development in the basal ganglia.^21,36^ This could be attributed to the impact of high pulse waves and restoration of damaged blood vessel walls, making the basal ganglia area more susceptible to damage than other regions. Furthermore, decreased elasticity and thickening of the vessel impair the contractile phenotype of smooth muscle cells, leading to reduced brain CBF.^4,30^ These could also cause alterations in the diameter and structure of small blood vessels, which in turn impairs fluid dynamics.^32^ All these structural changes ultimately lead to the expansion of the perivascular space, forming a pathological state.^33^

Alterations in the relationship between EPVS and CBF may be ascribed to regulating the glial-vascular unit (GVU), which comprises glial cells (astrocytes and microglia) and perivascular cells (endothelial cells, pericytes, and perivascular macrophages). The neurovascular unit (NVU) is widely acknowledged for its utility in investigating intricate interactions among multicellular structures.^37^ These cells collaborate to uphold central nervous system (CNS) homeostasis and execute diverse physiological functions. The interplay between glial cells and perivascular cells within the PVS microenvironment promotes the effective execution of multiple functions, including CBF regulation, angiogenesis, blood-brain barrier (BBB) integrity maintenance, and neurotoxic waste clearance through the glymphatic system.^38^ Some researchers have posited that GVU is a nexus for integrating neurovascular, neuroimmune, and neurodegenerative mechanisms after brain injury and during wound healing.^39^ Consequently, the heightened severity of EPVS impairs overall GVU function, leading to more pronounced changes in the cerebral vasculature, specifically reduced CBF, which is somewhat consistent with our results. Our findings indicated that the alteration in CBF was specifically linked to BG-EPVS rather than CSO-EPVS, thus implying that the damage caused by CSO-EPVS may be entering a compensatory phase or not primarily affecting the function of vascular component units, thereby allowing GVU’s regulation of CBF to remain unaffected. Nevertheless, further investigation is warranted.

In a study of patients with spontaneous intracerebral hemorrhage, the presence of severe CSO-EPVS served as an indicator of vascular amyloid burden that was helpful in the diagnosis of cerebral amyloid angiopathy, unlike BG-EPVS, thus suggesting that the spatial distribution of EPVS could reflect underlying cerebrovascular pathology.^40^ Abnormal protein aggregation, such as β-amyloid, can obstruct the upstream cortical artery system, impairing cerebrospinal fluid drainage and enlargement of the perivascular space.^8^ White matter exhibits lower cell density and greater susceptibility to stress than gray matter, which may contribute to the formation mechanism of CSO-EPVS.^4^ Two independent prospective cohort studies from the

United Kingdom and China, primarily including patients TIA or ischemic stroke, demonstrated that BG-EPVS, rather than CSO-EPVS, serve as prognostic markers for stroke and mortality, independent of other neuroimaging markers of CSVD.^25^ Our findings are consistent with this, indicating that the severity of CSO-EPVS is not significantly correlated with changes in CBF and that CSO-EPVS may not serve as an imaging marker for early ischemic stroke.

There are relatively few risk factors associated with EPVS. A previous study demonstrated that age, hypertension, and WMH were the primary influencing factors of BG-EPVS,^41^ which is in line with our findings. Some studies have also indicated that in addition to gender, age, obesity, and other influencing factors, EPVS was associated with cognitive dysfunction and WMH.^19^ Our results revealed that hypertension, drinking, sleep duration, and PVWMH were all correlated with CSO- EPVS, contradicting some existing literature. For example, Yamasaki et al. identified no specific risk factors related to CSO-EPVS, possibly due to differences in the included population and an age limit of 40 years old.^38^ Previous research has shown that EPVS could lead to microdamage in the white matter of the brain, which is closely tied to WMH. EPVS and WMH may share common pathogenic mechanisms associated with interstitial fluid leakage, microvascular wall dysfunction, and vascular inflammation.^39,40^ While it has been reported that neuronal atrophy may cause EPVS in the basal ganglia, we found no correlation between EPVS and brain volume. Current research results present inconsistencies requiring further investigation.^9^ Other studies have suggested associations between EPVS, breathing patterns, sleep quality, and weight management.^42^ The current literature reports different results on the risk factors related to EPVS; in short, there is a possibility that a combination of hitherto unknown environmental and genetic factors may be at play.^43^

The present study has several limitations. Firstly, as this is a single-center study with a limited sample size, future research should focus on extending to multiple centers rather than just increasing the sample size. Secondly, the brain CBF exhibits continuous fluctuations despite our efforts to maintain subjects’ calmness. Consequently, it is challenging to obtain completely accurate and consistent measurements of blood flow. Thirdly, setting the post-label delay (PLD) parameters in the ASL sequence may have specific effects on quantitative CBF values. Also, when choosing the region of interest (ROI), it is unclear whether alternative options, such as functional partitioning, would yield meaningful results based on the analysis of brain anatomical structure.

## Conclusion

Our data indicated that BG-EPVS, rather than CSO-EPVS, was negatively correlated with CBF across the whole brain and almost all brain regions. Also, CBF decreased with the increasing severity of BG-EPVS, suggesting that BG-EPVS could serve as an imaging marker for reflecting the changes of brain CBF and an effective indicator for early ischemic stroke.

## Data Availability

Data availability statement: All relevant data are within the paper.

## Acknowledgments

We are deeply grateful to our patients and their families for their enthusiastic participation in our study and to the staff of Longgang Central Hospital for their assistance in conducting medical interviews and obtaining clinical data.

## Abbreviations

EPVS: enlarged perivascular spaces
MRI: magnetic resonance imaging
BG: basal ganglia
CSO: centrum semiovale
CBF: cerebral blood flow
ASL: arterial spin labeling
CSVD: cerebral small vessel disease
WMH: white matter hyperintensities
TIA: transient ischemic attack
GVU: glial vascular unit
GMV: gray matter volume,
WMV: white matter volume
CSF: cerebrospinal fluid

